# Comparison between mid-nasal swabs and buccal swabs for SARS-CoV-2 detection in mild COVID-19 patients

**DOI:** 10.1101/2022.01.20.22269539

**Authors:** Ignacio Blanco, Concepción Violán, Clara Suñer, Julio Garcia-Prieto, Maria José Argerich, Meritxell Rodriguez-Illana, Nemesio Moreno, Pere-Joan Cardona, Anna Blanco, Pere Torán-Monserrat, Bonaventura Clotet, Josep M^a^ Bonet, Nuria Prat

## Abstract

**Background:** The use of rapid antigen diagnostics tests (Ag-RDT) has gained widespread acceptance as an alternative method for diagnosis of COVID-19 outside of health care settings. Various authors have reported that saliva is a reliable specimen, alternative to nasopharyngeal and mid-nasal swabs, to detect SARS-CoV-2 infections by RT-PCR. We assessed the performance of buccal swabs containing saliva for SARS-CoV-2 detection by Ag-RDT, using mid-nasal specimens as a reference in the northern area of Barcelona (Catalonia, Spain)

**Methods:** In the context of routine clinical diagnosis of mild COVID-19 patients, we enrolled 300 adults in a study to directly compare mid-nasal swabs and saliva specimens for SARS-CoV-2 detection by Ag-RDT.. When mid-nasal and buccal Ag-RDTs showed discordant results, a third mid-nasal swab was collected and analysed by RT-PCR.

**Results:** Paired samples were successfully obtained in 300 suspected cases of SARS-CoV-2 infection. Of the 300 paired samples, Ag-RDT with the mid-nasal swab detected 139 (46.3%) positive COVID-19 cases. In comparison, buccal swabs showed a sensitivity and specificity of 31.7% (44/139) and 98.8% (159/161), respectively. 65 discordant results with positive mid-nasal swabs and negative buccal swabs were tested by RT-qPCR. All samples tested by Rt-PCR resulted positive, with a mean cycle threshold (Ct) of 28.3 (SD 7.3).

**Conclusion:** Our findings show that mid-nasal swabs have better performance than buccal swabs for detecting SARS-CoV-2 with Ag-RDT tests. Of note, the sensitivity of buccal samples was affected in samples with high viral loads (Ct<33), suggesting that buccal swabs might not be sensitive enough to detect individuals at risk of transmission. Taken together, the existing literature and the results provided in our analysis we advise against the use of buccal specimens for SARS-CoV-2 diagnostics with Ag-RDT.

## Background

The use of rapid antigen diagnostics tests (Ag-RDT) has gained widespread acceptance as an alternative method for diagnosis of COVID-19 outside of health care settings. Ag-RDT offer advantages as they can be deployed by members of the general public, which require the use of self-collected specimens. Various authors have reported that saliva is a reliable specimen, alternative to nasopharyngeal and mid-nasal swabs, to detect SARS-CoV-2 infections by RT-PCR.^1–4^ Regarding the use of Ag-RDTs with saliva samples, previous studies have mainly reported limitations on the ability of Ag-RDT for COVID-19 diagnosis in this specimen.^5,6^ These limitations could be derived on the viral load distribution or sample preparation protocols, which might need to be adapted to the rheological properties of saliva. Therefore, even if several commercialized Ag-RDT tests list saliva as a possible specimen, the European Centre for Disease Prevention and Control (ECDC) currently only validates tests based on nasal, oropharyngeal and/or nasopharyngeal specimens.^7^

SARS-CoV-2 variants are characterized by distinct mutations, which impact on disease transmissibility, immune escape, diagnostics and possibly tissue tropism. A preliminary study has proposed that saliva swabs are the preferred sample for Omicron variant detection by RT-PCR.^8^ Therefore, during a surge of the Omicron variant in the northern area of Barcelona (Catalonia, Spain), we assessed the performance of buccal swabs containing saliva for SARS-CoV-2 detection by Ag-RDT, using mid-nasal specimens as a reference.

## Methods

In the context of routine clinical diagnosis of mild COVID-19 patients, we enrolled 300 adults in a study to directly compare mid-nasal swabs and saliva specimens for SARS-CoV-2 detection by Ag-RDT. Participants should not have had any food, drink, tobacco or gum in the 30 minutes preceding saliva swab collection. Participants were initially instructed to cough 3-5 times, wearing a surgical mask. Buccal and mid-nasal swabs were collected by health workers, and mid-nasal swabs were used as a reference. Each swab specimen was then used for Ag-RDT detection following manufacturer’s instructions (tests used: BIOSYNEX COVID-19 Ag BSS (ref SW40006); FLOWFLEX™ (ref L031-11815); Panbio COVID-19 Ag Test (Abbott)). When mid-nasal and buccal Ag-RDTs showed discordant results, if the individual consented, a third mid-nasal swab was collected and analysed by RT-PCR. This third swab specimen was placed into a sterile tube containing viral transport media (DeltaSwab Virus) and transported to the Microbiology laboratory of *Hospital Germans Trias i Pujol* and stored at 2 – 8 ºC for up to 24 hours before RT-qPCR. RNA was extracted using the STAR Mag reagent (Seegen) for the Microlab Starlet IV or Nimbus platforms (Hamilton life Science Robotics, USA), according to the manufacturer’s instructions. PCR amplification was conducted according to the recommendations of the 2019-nCoV RT-qPCR Diagnostic Panel of the Centers for Disease Control and Prevention (CDC) using the Allplex™ 2019-nCoV assay (Seegene, South Korea) on the CFX96 (Bio-Rad, USA) according to manufacturer’s instruction.

## Results

Paired samples were successfully obtained in 300 suspected cases of SARS-CoV-2 infection. Included participants had a mean age of 43.6 years (SD 14.6) and 59.7% were females. 285 (95.0%) participants were symptomatic. The mean number of symptoms was 3.6 (SD 1.8) and the median time from symptom onset was 2 days (IQR 1-3). The most frequently reported symptoms were headache (65.7%); sore throat (64.3%); cough (61.0%) and rhinorrhoea (60.0%).

Of the 300 paired samples, Ag-RDT with the mid-nasal swab detected 139 (46.3%) positive COVID-19 cases. In comparison, buccal swabs showed a sensitivity and specificity of 31.7% (44/139) and 98.8% (159/161), respectively (Figure 1). 65 discordant results with positive mid-nasal swabs and negative buccal swabs were tested by RT-qPCR. All samples tested by Rt-PCR resulted positive, with a mean cycle threshold (Ct) of 28.3 (SD 7.3).

**Figure 1.**
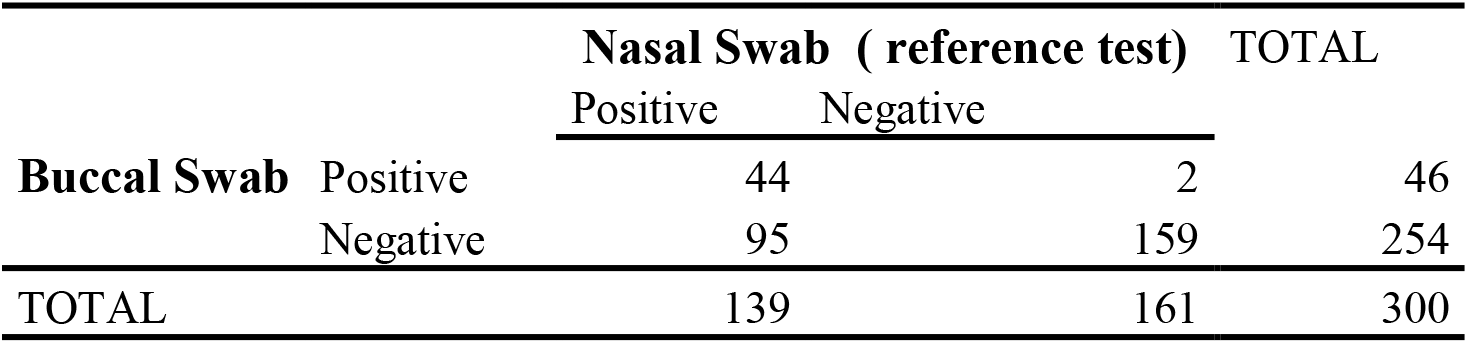
Diagnostic accuracy of buccal swab-Ag-RDT versus standard Ag-RDT on nasal swab.

## Discussion

In summary, our findings show that mid-nasal swabs have better performance than buccal swabs for detecting SARS-CoV-2 with Ag-RDT tests. Of note, the sensitivity of buccal samples was affected in samples with high viral loads (Ct<33)^9^, suggesting that buccal swabs are not sensitive enough to detect individuals at risk of transmission. Regarding the usability of buccal swab samples, due to the previous coughing step and the time required for its collection, they imply a higher exposure of the health worker to the potentially infected person.

Taken together, the existing literature and the results provided in our analysis we advise against the use of buccal specimens for SARS-CoV-2 diagnostics with Ag-RDT.

## Data Availability

All data produced in the present study are available upon reasonable request to the authors

## Acknowledgments

We thank the North Metropolitan Management for the support in the development of the project. We thank the personnel from the Centralized Support Unit COVID-19 (UCSC), Primary care emergency centres and Northern Metropolitan Clinic Laboratories for their support in administration, human resources, and supply chain management. The authors declare no conflicts of interest.

## Ethics

The study was conducted according to the Helsinki Declaration of the World Medical Association. The study protocol was approved by the Ethics Committee at IDIAP Jordi Gol and the institutional review boards of participating centres (P22/022). All patients provided written informed consent before enrolling the study, which was supervised by an independent data and safety monitoring board.

## Data sharing

All data produced in the present study are available upon reasonable request to the authors.

